# Enhanced recovery after surgery (ERAS) protocols is extremely beneficial in liver surgeries – A metaanalysis

**DOI:** 10.1101/2020.04.11.20061689

**Authors:** Bhavin Vasavada, Hardik Patel

## Abstract

**BACKGROUND:** Enhanced recovery after surgery (ERAS) programs aim to improve postoperative outcomes.. This metaanalysis aims to evaluate the impact of ERAS programmes on outcomes following liver surgeries.

**METHODS:** EMBASE, MEDLINE, PubMed and the Cochrane Database were searched for studies comparing outcomes in patients undergoing liver surgery utilizing ERAS principles with those patients receiving conventional care. The primary outcome was occurrence of 30 day morbidity and mortality. Secondary outcomes included length of stay, functional recovery, readmission rates,time to pass flatus,blood loss and hospital costs.

**RESULTS:** Ten articles were included in the metaanalysis. 30 days morbidity and mortality was significantly less in ERAS group.Hospital stay, time to pass flatus, time to complete recovery and hospital costs were also significantly reduced due to ERAS protocols. Blood loss and readmission rates were also significantly less in ERAS group.

**CONCLUSIONS:** The adoption of ERAS protocols significantly reduced morbidity, mortality hospital stay, readmission rates, time to recovery, hospital costs, time to pass flatus, blood loss and readmission rates.

## Introduction

Early recovery after surgery (ERAS) protocol is beccoming gold standard in peri operative care with excellent reults in colorectal,gastiric and HPB surgeries. [1].

ERAS is a evidence based peri-operative protocol which has shown significnat improvements in perioperative outcomes. [2]. Despite these overwhelming evidences implementation of these protocols has been very slow and lack wide spread implementation.[3]

ERAS has initially developed for colorectal surgeries [4], However its implementation is being tested in all other field.[4] and it has now spread over other specialities.

ERAS protocols has been applied to liver surgeries also and found to be benefical.[5]

Primary Aim of this metaanalysis was to study the effect of ERAS protocols on 30 days morbidity and mortality. Secondary aim was to study effect of ERAS protocols on hospital stay, readmission rates, time to recovery, time to pass flatus, and Hospital costs.

## Material and Methods

In this systemic review and metaanalysis we searched EMBASE, MEDLINE, PubMed and the Cochrane Database with key words like “liver surgery”,”Enhanced recovery after surgery”, “ERAS protocols”, “ERAS vs conventional liver surgery”, “ morbidity and mortality following liver surgery”, ‘liver resections”. Two independent authors extracted the data (B.V and H.P).

Systemic review and Metaanalysis was done according to MOOSE and PRISMA guidelines. (6,7).

## Statistical analysis

The meta-analysis was conducted using Open metaanalysis software. Heterogeneity was measured using Q tests and I^2^, and p < 0.10 was determined as significant (8). If there was no or low heterogeneity (I < 25 %), then the fixed-effects model was used. Otherwise, the random-effects model was used. The risk ratio (RR) was calculated for dichotomous data, and weighted mean differences (WMD) were used for continuous variables. Both differences were presented with 95 % CI. For continuous variables, if data were presented with medians and ranges, then we calculated the means and SDs according to Hozo et al. (9). If the study presented the median and inter-quartile range, the median was treated as the mean, and the interquartile ranges were calculated using 1.35 SDs, as described in the Cochrane handbook.

## Inclusion criteria

**Inclusion criteria:**

1. Studies that compared ERAS protocols with that of conventional protocol
2. Minimum 25 numbers of patients
3. Means and standard devations or medians and range mentioned.
4. Full texts available
5. Prospective, retrospectives studies or randomised control trials included.
6. ERAS program should include most of the 17 items included according to ERAS group recommendation. [10].

### Exclusion criteria

1. Studies whose full texts can not be obtained.
2. Studies with no comparable groups [ERAS vs conventional]
3. Duplicate studies.

## Assessment of Bias

Characteristics of the studies are described in table 1. Identified studies were broadly grouped into 1 of 2 types, either randomized trials or cohort studies. Cohort studies were assessed for bias using the Newcastle-Ottawa Scale ^(10)^. Randomized trials were assessed based on the Cochrane Handbook. (11) (Table 2 and table 3)

**Table 1.** charecteristics of studies.

| Study | Type of study | Number of patients<br>in ERAS group | Number of patients<br>in control group |
| --- | --- | --- | --- |
| bobbyv2015 | COHORT | 91 | 93 |
| vandam2008 | COHORT | 61 | 100 |
| koea2009 | COHORT | 50 | 50 |
| lin2011 | COHORT | 56 | 61 |
| jones2013 | RCT | 46 | 45 |
| ni2013 | RCT | 80 | 80 |
| sanchez2012 | COHORT | 26 | 17 |
| HeF2015 | RCT | 48 | 38 |
| lu2014 | RCT | 80 | 80 |
| liang 2016 | RCT | 80 | 107 |

**Table 2:** Risk of bias summary of RCT. + denotes low risk of bias, – denotes high risk of bias.

|  | Random Sequence generation | Allocation Concealment | Performance Bias | Detection Bias | Attrition Bias | Reporting Bias | Other |
| --- | --- | --- | --- | --- | --- | --- | --- |
| jones2013 | + | + | - | + | + | + | ? |
| ni2013 | + | + | - | - | + | + | + |
| HeF2015 | + | + | - | + | + | + | + |
| lu2014 | ? | ? | - | + | + | + | ? |
| liang2016 | ? | ? | - | - | + | + | - |

**Table 3.** Assessment of bias in cohort studies. + Denotes low risk of bias, - denotes high risk of bias.

|  | Representative of exposed cohort | Selection of non exposed cohort | Ascertainment of Exposure | Demonstration that outcome was not present at start of study | Comparability of cohorts | Assessment of outcomes | Adequate time for followup | Complete Follow up of cohort | Total score |
| --- | --- | --- | --- | --- | --- | --- | --- | --- | --- |
| bobbyv2015 | + | + | + | + | + | - | + | + | 7 |
| vandam2008 | + | + | + | + | - | - | + | + | 6 |
| koea2009 | + | - | - | + | - | - | + | + | 4 |
| lin2011 | + | + | + | + | + | - | + | + | 7 |
| sanchez2012 | + | + | + | + | - | - | + | + | 6 |

## Results

### Search results

Total 190 studies identified from initial literature search, 157 studies were evaluated after duplicates removed. Only 57 studies included ERAS protocols, 34 studies full text obtained. 13 studies had comparable groups for conventional protocols. Out of it 10 studies included in final analysis as other studies did not include adequate ERAS protocols. [figure 1]. (13–22)

**Figure 1.**
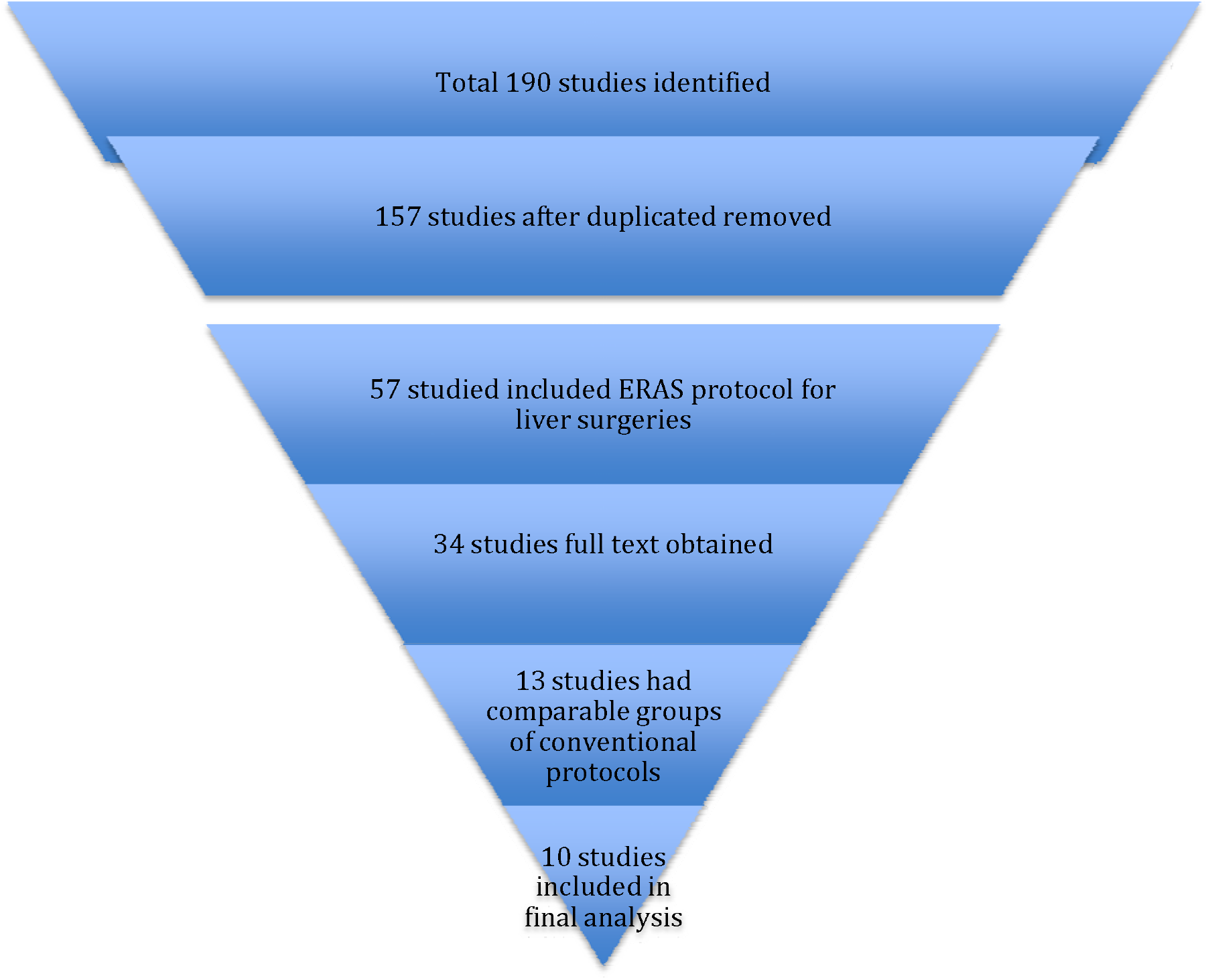
Search strategy according to PRISMA guidelines.

**Figure 2.**
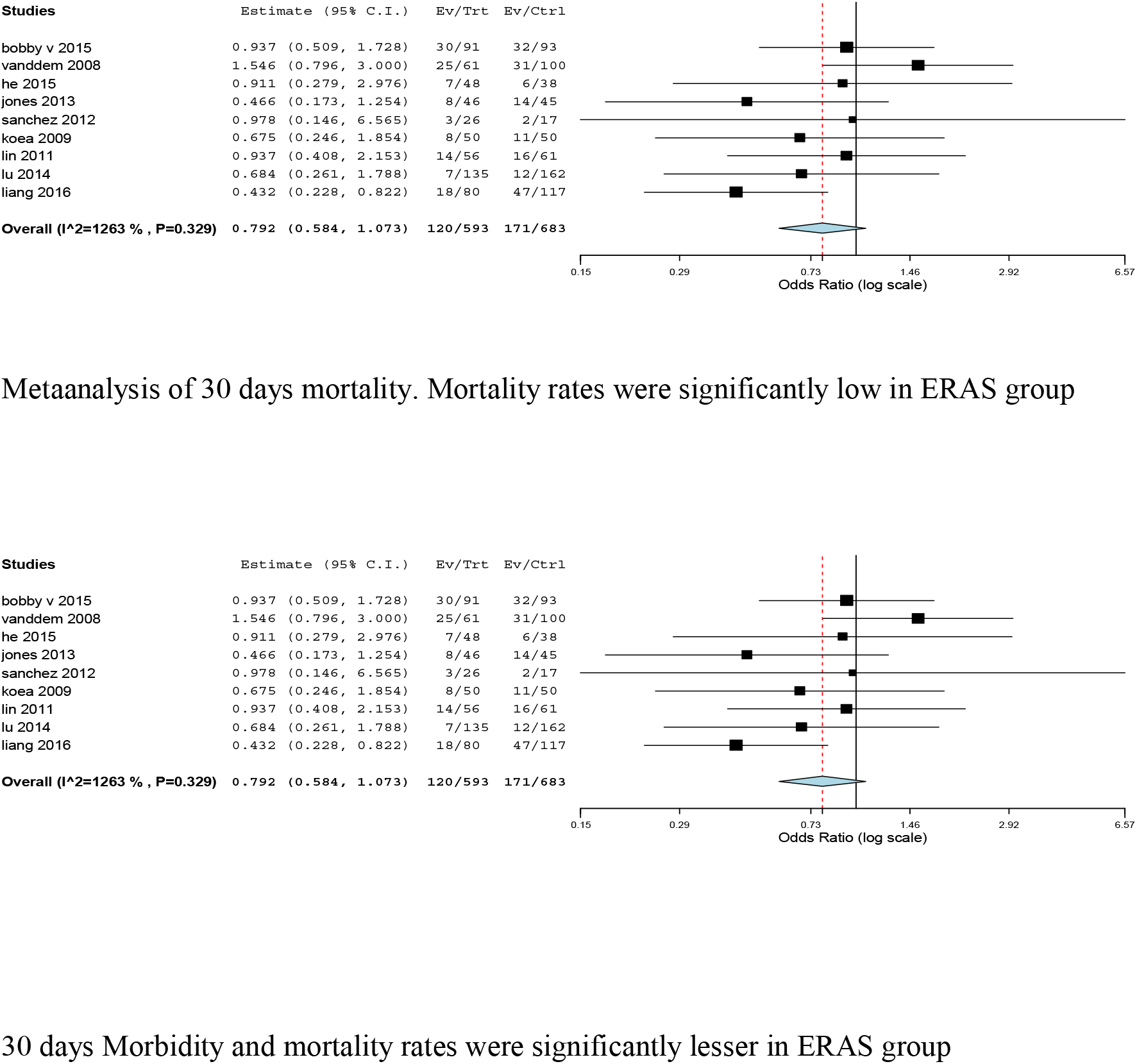
metaanalysis of 30 days mortality and morbidity rates between ERAS vs conventional approach.

Total 1289 patients’ outcomes were studied from these 10 studies. 618 in ERAS group and 618 in conventional group.

## Metaanalysis

### Primary outcome measures

30 days mortality:

3 patients died in ERAS out of 458 and 5 patient died in conventional approach out of 511. Mortality was significantly less (p =0.029)

30 days morbidity:

30 days morbidity rates were significantly less. P <0.001.114/593 patients developed complications in ERAS group vs 171/673 in conventional group.

## Secondary outcomes

We also evaluated hospital stay,time to functional recovery, readmission rates, time to pass flatus, hospital coses and blood loss in ERAS protocols in liver surgery.

As shown in figure 3 hospital stay (p<0.001 WMD −2.191 and time to functional recovery (p<0.001, WMD −2.462) were significantly less in ERAS group. Readmission rates were also significantly less in ERAS group.

**Figure 3.**
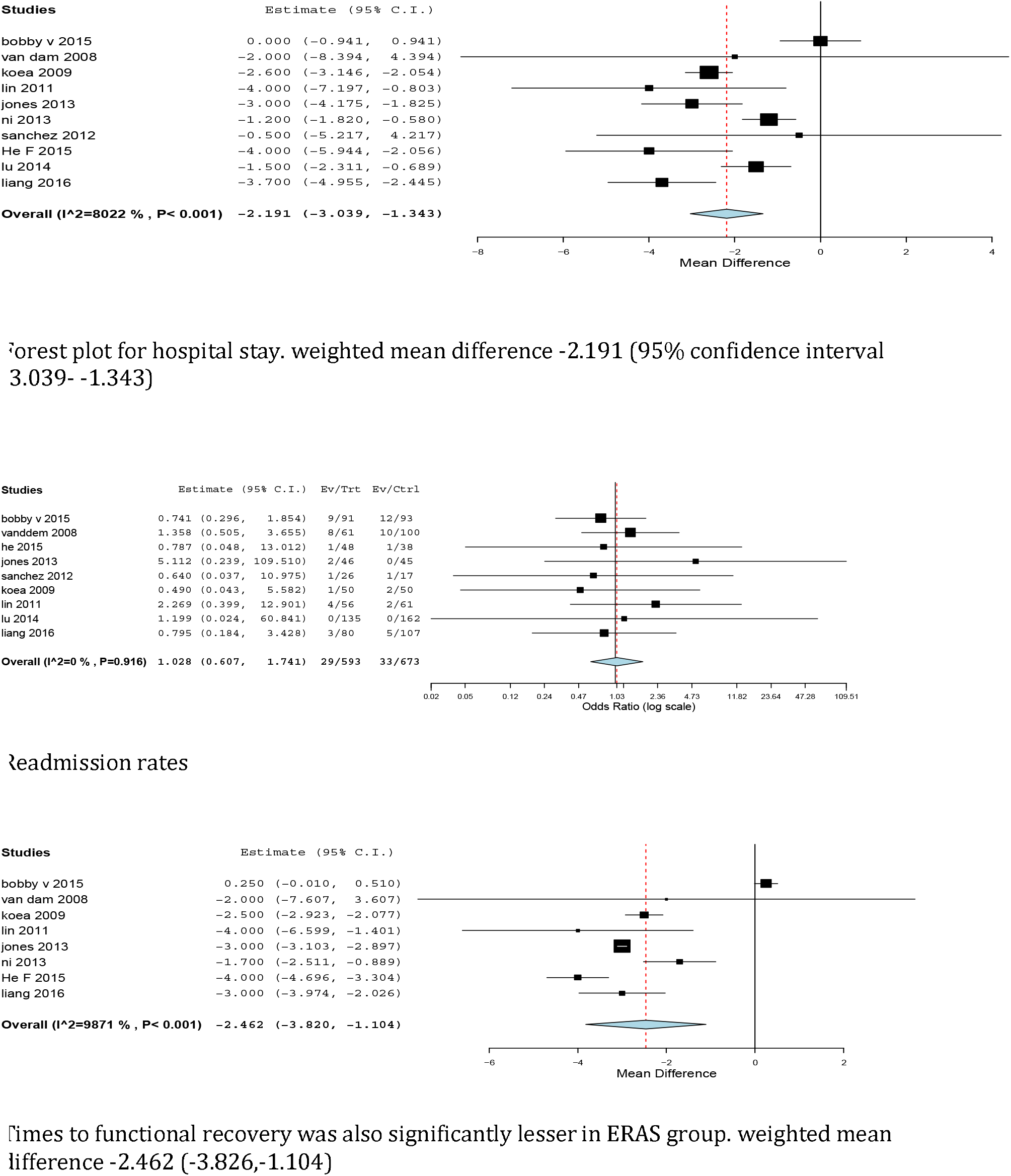
metaanalysis of hospital stay, readmission rates and time to functional recovery

There was significantly less blood loss in ERAS group. (p<0.001) (figure 4). Time to pass flatus and hospital costs were significantly lesser in ERAS group. (p= 0.035 and p <0.001 respectively with WMD of −0.996 days and – 1803.536 $ respectively).

**Figure 4.**
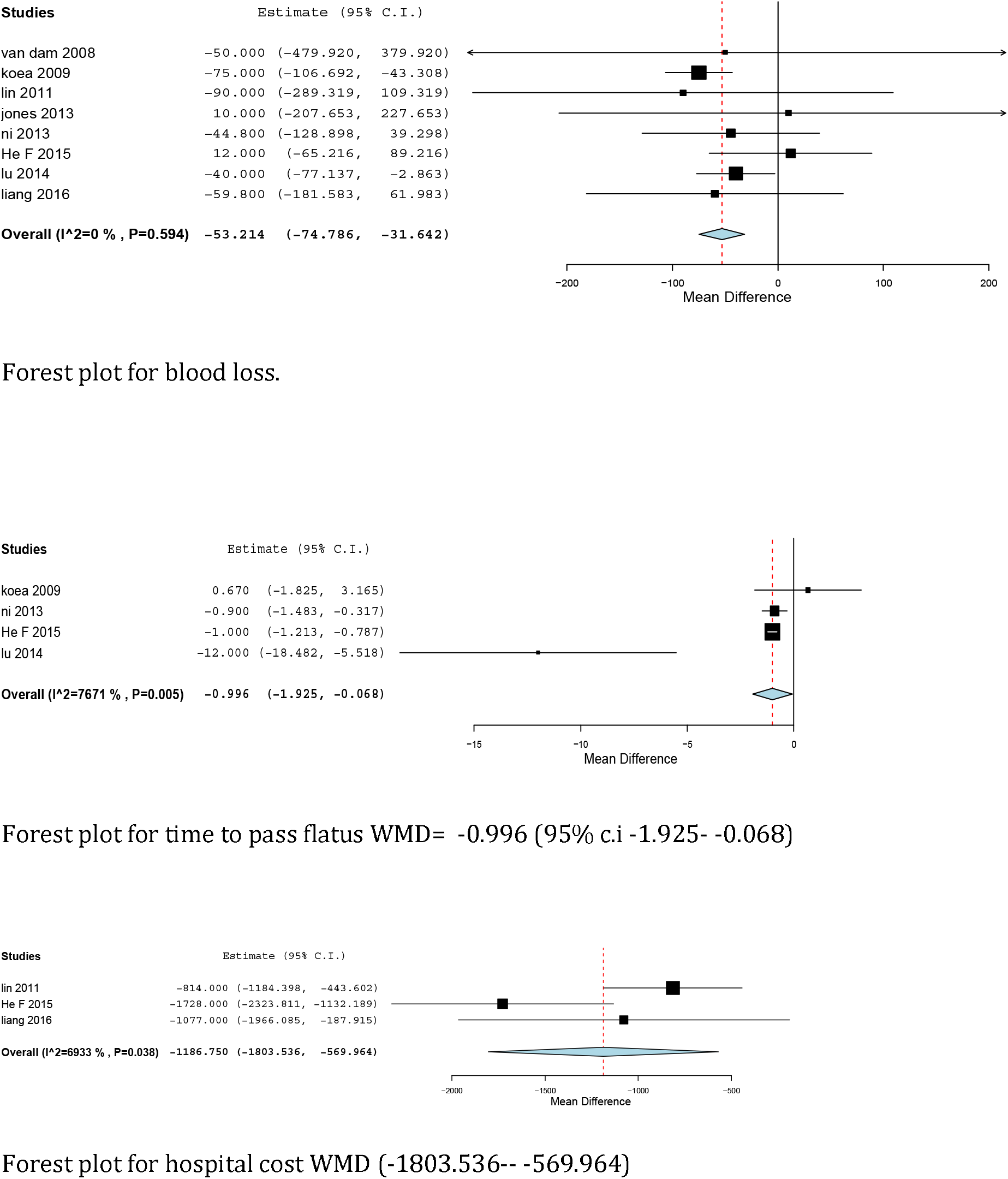
Metaanalysis for blood loss, time to pass flatus and hospital cost

## Discussion

Enhanced recovery after surgery though initially described for colorectal surgery is now becoming standard protocol for all surgeries and it has significantly reduced hospital stay and cost without affecting morbidity and mortality.[1–5]

Started from colorectal surgeries ERAS protocols has now moved to other branches of surgeries. Many authors have tried to study applications of ERAS protocols on liver surgeries. (13–22) and showed ERAS protocol has significant benefit over standard protocols however large number studies and quality metaanalysis are still missing. Purpose of this metaanalysis to compare outcomes between ERAS and conventional group.

After literature review we evaluated 10 studies in this metaanalysis 4 were Randomised cotrol trials (11–14) and 6 were prospective or retrospective cohort studies. (15–20).

We evaluated 30 days mortality and morbidity as primary outcomes and hospital stay,time to complete recovery (time to complete physical independence), readmission rates,time to pass flatus, blood loss and hospital costs as secondary out comes.

There was significantly less mortality and morbidity in ERAS group. (figure 2). Hospital stay, time to functional recovery and time to pass flatus (4 studies) were also significantly different in both the groups. (WMD −2.191,Odds ratio 0.016, and WMD-2.462 respectively).

Blood loss and readmission rate was significantly less in ERAS group.. Only 3 studies out of 10 evaluated hospital cost which was signiicantly lesser in ERAS group. (WMD −1803.536$).

There are some limitations of this metaanalysis as heterogeneity of studies was significantly random effect models were used. Except hospital stay at least one study did not evaluate other factors.

In conclusion ERAS programs in liver surgeries reduces morbidity, mortality hospital stay, readmission rates, time to recovery, time to pass flatus, hospital cost and blood loss and it is extremely beneficial in liver surgeries.

## Data Availability

data will be made available on demand

**Abbreviations**
Enhanced Recovery After Surgery: (ERAS)
Weighted Mean Difference: (WMD)
Confidence Intervals: (C.I)

## Contribution of authors

Dr.bhavin vasavada was over all incharge of research and actively contrubuted in study design,data collection, conduction research,statistics, manuscript written, final approval. Dr.hardik patel helped in data collection, conduction research,statistics, manuscript writing.

